# Screening Outcomes and Associated Factors in a Follow-up Cervical Cancer Screening among Women Living with HIV, 2019–2024: Evidence from Moshi Municipality

**DOI:** 10.1101/2025.11.26.25341052

**Authors:** Dickson Doto, Laura Shirima, Blandina Mmbaga, Deogratius Kinoko, Innocent Mboya, Innocent H. Peter Ughh, Alex Mremi, Patricia Swai, Bariki Mchome

## Abstract

**Background:** Cervical cancer screening is a critical public health intervention that facilitate the early detection and treatment of cervical precancerous lesions before they progress to invasive cancer. The effectiveness of this intervention largely depends on timely and consistent adherence to recommended follow-up screenings. However, in Tanzania, the screening outcomes in follow-up screenings among WLHIV and its associated factors remain poorly understood.

**Objective:** To determine the screening outcomes in a follow-up screening and its associated factors among WLHIV who have undertaken their first screening between 2019 and 2022 in Moshi Municipality.

**Methodology:** This was a retrospective cohort study. Data were analyzed using STATA version 18.0. Descriptive statistics were employed to determine the screening outcomes in a follow-up screening. Multivariable logistic regression analysis was used to estimate adjusted odds ratios and 95% CI for the factors associated with the screening outcomes.

**Results:** Among 1,019 WLHIV who adhered to a follow-up screening, only 43(4.2%) were diagnosed with abnormal screening outcome. Majority, 38(88.4%) of those diagnosed with abnormal screening outcomes were also previously diagnosed with such screening outcomes during their first screening and received treatment. Moreover, WLHIV with abnormal screening outcomes in their first screening were significantly more likely to have abnormal screening outcome again during their follow-up screening compared to those who initially screened normal [AOR=12.51, 95%Cl: 8.92 – 37.64].

**Conclusion:** Abnormal screening outcomes in a follow-up screening was low. Improvements in the providing treatment for cervical precancerous lesions after the diagnosis in the first screening is essential to further minimize the proportion of abnormal screening outcomes in follow-up visit

## Introduction

Cervical cancer screening is the crucial strategy in the fight against cervical cancer (1). It allows for the early detection and treatment of precancerous cervical lesions before they progress into invasive cervical cancer (2). Numerous studies have demonstrated that early cervical cancer screening combined with timely follow-up screenings can significantly reduce the burden of the disease (3,4). For instance, follow-up screening every 10 years can reduce the burden of cervical cancer by 40-60%, every 5 years by 85%, every 3 years by 91% and annually by 94% (5).

Much of the global disease burden is borne in sub-Saharan Africa where approximately 85% of cervical cancer cases and 90% of related deaths occurs (6). The disease is also much more prevalent among WLHIV compared to the women in the general population (7,8). In response to this increased risk, WLHIV in Tanzania are eligible for their first cervical cancer screening as early as 25 years of age, compared to 30 years for women without HIV (9). Also, WLHIV are recommended to undertake a follow-up screening after a year compared to 3 years recommended for women with no HIV. Moreover, Visual Inspection with Acetic Acid (VIA) is currently the recommended cervical cancer screening method for WLHIV in the country. However, HPV DNA test is under pilot implementation and is expected to be incorporated soon as an optional screening method (10).

Studies across the globe have indicates that women continue to present with abnormal screening outcomes in a follow-up screening despite having been treated for cervical precancerous lesions detected during their first screening (11). However, there is existence of disparity particularly between developed and underdeveloped countries. However, notable disparities exist between developed and developing countries (12–15). To our knowledge, no study in Tanzania has specifically examined this issue, Therefore, this study aimed to determine follow-up screening outcomes and their associated factors among WLHIV, using Moshi Municipality as the study area.

## Materials and Methods

This was a retrospective cohort study, aiming to determine the screening outcomes in a follow-up screening and its associated factors among WLHIV who had undertaken their first screening between 2019 and 2022 in Moshi Municipality. For cervical cancer screening data, the study utilized data recorded in the National Health Management Information System (HMIS), which is locally known as *Mfumo wa Taarifa za Uendeshaji wa Huduma za Afya* (MTUHA). This system comprises both electronic and hardcopy registries. The electronic component contains aggregated summaries while the hardcopy registries contain detailed, individual-level screening records. The system is designed to ensures consistency and standardization in data collection and reporting, thereby providing a reliable source of information for monitoring and evaluating the cervical cancer screening programme in the country (16). Meanwhile for HIV related data, the study utilized CTC2 database, and the information of the study participants in that database were linked with their information in the cervical cancer screening registries, using their unique identification numbers.

Moreover, four healthcare facilities were purposively selected out of six which offer cervical cancer screening services in the Municipality. This includes Mawenzi Regional Referral Hospital, St. Joseph Hospital, Majengo Health Centre and *Chama cha Uzazi na Malezi Bora* (UMATI). The study area was purposively selected because the Kilimanjaro region, which the Moshi Municipality is the part of, has consistently been ranked as the region with the highest prevalence of cervical cancer in the country (17). Additionally, the region is among the leading regions in the country in terms of the number of women screened for cervical cancer. So the selection of this study area ensured the inclusion of the most affected region by the disease in the country and guaranteed the availability of sufficient and reliable data to effectively address the study objectives (18).

Descriptive statistics were used to summarize the socio-demographic and clinical characteristics of the study participants. The results for categorical variables were presented using frequencies and proportions, while results for continuous variables were summarized using means and standard deviations. Moreover, STATA 18.0 software was used for data cleaning and analysis. The difference in the screening outcomes in a follow-up screening by independent variables was assessed using the Chi-square test or Fisher’s exact test (when expected cell counts were less than 5). Moreover, multivariable logistic regression with robust standard errors was used to determine the odds ratios (OR) and their corresponding 95% confidence intervals (CI) for the factors associated with the screening outcomes in a follow-up screening.

Moreover, prior to model fitting, key assumptions of logistic regression were thoroughly assessed. To assess for the presence of outliers in the continuous variables - age and parity, boxplot diagrams were used. When evidence for outliers was detected, the winsorization technique was applied to remove the outliers from those variables, and as a result, new variable without outliers were then generated, categorized into distinct groups based on the previous studies, and subsequently used in the model fitting (19). Also, to assess for the presence of multicollinearity among the independent variables, the spearman correlation technique was employed. In cases where multicollinearity was evident, one of the highly correlated variables was excluded. Moreover, alternative to logistic regression models were not used for this case because the prevalence for cervical cancer screening in the Municipality was above 10%, which is the cut-off point for opting for such models.

Moreover, in the model building, crude odds ratio (COR) was first estimated for each independent variable. Variables found to be statistically significant at p-value <0.05, those with potential confounding effect and known clinical association with the screening outcomes in a follow-up screening were included in the multivariable logistic regression model. Additionally, potential interaction effects among the independent variables were also assessed during the multivariable analysis. However, evidence for such effect was not found. Also, both forward and backward stepwise logistic regression techniques were used in the multivariable analysis to identify the most parsimonious model and estimate adjusted odds ratio (AOR) for the predictors. Moreover, model selection was guided by the Akaike Information Criteria (AIC), whereby the model with the lowest AIC value was selected as the final model.

## Results

A total of 3,076 participants were included in the study. Their mean age at the time of their first screening was 39.3 years, with the highest proportion, 1,121(36.4%), aged between 35 and 44 years. Most participants, 3,058(99.4%), were residing in Kilimanjaro region. Most participants, 2,557(83.1%), received their first screening at the healthcare facility rather than at an outreach facility. Additionally, the mean parity among the study participants was 2.7, with majority proportion, 2,213(71.9%), had between 2 and 4 parities at the time of the first screening.

Also, majority, 2,771(90.1%), had no other reproductive comorbidities. Most, 973(31.6%), were classified as having WHO HIV stage I at the time of the first screening. Moreover, regarding HIV viral load, majority, 2,360(76.7%), had undetectable levels (*<*50 copies/mm^3^) at the time of the first screening. Also, majority, 2,201(71.6%), had not yet developed Advanced HIV Disease (AHD) at the time of the first screening. Additionally, all of the screening procedures were performed by VIA method. Notably, all healthcare providers involved in the first screening, treatment of precancerous cervical lesions and follow-up screening were females **(Table 1)**.

**Table 1:** Socio-demographic and clinical characteristics of the study participants (n=3,076)

| **Characteristics** | **n(%)** |
| --- | --- |
| **Age (years)** |  |
| *Mean (Standard deviation)* | 39.3(9.3) |
| <25 | 177(5.8) |
| 25-34 | 818(26.6) |
| 35-44 | 1,121(36.4) |
| 45-54 | 848(27.6) |
| ≥55 | 112(3.6) |
| **Area of residence** |  |
| Residing in Kilimanjaro region | 3,058(99.4) |
| Residing in other regions | 18(0.6) |
| **Screening location** |  |
| Healthcare facility | 2,557(83.1) |
| Outreach facility | 519(16.9) |
| **Healthcare facility involved** |  |
| Majengo Health Centre | 1,025(33.3) |
| Mawenzi Regional Referral Hospital | 1,537(50.0) |
| St. Joseph Hospital | 472(15.3) |
| UMATI | 42(1.4) |
| **Parity at the first screening** |  |
| *Mean (Standard deviation)* | 2.7(1.4) |
| 0-1 | 582(18.9) |
| 2-4 | 2,213(71.9) |
| ≥ 5 | 281(9.2) |
| **Information source for HIV status of the patients** |  |
| Self-declared | 312(10.2) |
| CTC card | 2,520(81.9) |
| PICT | 215(7.0) |
| RCH4 card | 10(0.3) |
| RCH5 card | 19(0.6) |
| **Presence of other reproductive comorbidities at the first screening** |  |
| No | 2,771(90.1) |
| Yes | 305(9.9) |
| **WHO HIV stage of the patients at the first screening\*** |  |
| I | 973(31.6) |
| II | 441(14.3) |
| III | 559(18.2) |
| IV | 177(5.8) |
| **HIV viral load of the patients at the first screening(copies/mL) \*** |  |
| Undetected (<50) | 2,360(76.7) |
| Suppressed (50-1000) | 91(2.9) |
| Unsuppressed (>1000) | 47(1.5) |
| **Advanced HIV Disease (AHD) status of the patients at the first screening\*** |  |
| No | 2,201(71.6) |
| Yes | 35(1.1) |

**Key:** n (%) do not tally to 100% due to missing values, **CTC card:** Care and Treatment Clinic card people living with HIV, **PICT:** Provider-Initiated HIV Counselling and testing card, **RCH4 card:** Reproductive and Child Health Card for Pregnant Women, **RCH5 card:** Reproductive and Child Health Card for Family Planning Services.

### Bivariate and multivariate analysis

Among 1,019 WLHIV who adhered to a follow-up screening, only 43(4.2%) [95%CI: 3.07% – 5.64%] were diagnosed with abnormal screening outcome whereas 976(95.8%) [95%CI: 94.3% – 96.9%] were diagnosed with normal screening outcome. All of the abnormal screening cases were categorized as small precancerous cervical lesions. Additionally, 38(88.4%) [95%CI: 74.9% – 96.1%] of those diagnosed with abnormal screening outcomes in a follow-up screening were also previously diagnosed with such screening outcomes in a first screening and received treatment for precancerous cervical lesions thereafter while the remaining 5(11.6%) [95%CI: 3.8% – 25.1%] were diagnosed with normal screening outcomes.

343(35.2%) of the participants who went to a referral healthcare facility for the first screening were diagnosed with normal screening outcomes in a follow-up screening. 329(3.3%) of the participants who were aged below 25 years at the time of the first screening were diagnosed with normal screening outcome in a follow-up screening. Also, 341(43.9%) of the participants who were at WHO HIV stage 1 at the time of the first screening were diagnosed with normal screening outcome in a follow-up screening. Also, 916(93.9%) of the participants diagnosed with normal screening outcome at the time of the first screening were again diagnosed with normal screening outcome in a follow-up screening. Additionally, 19(37.3%) of the participants who were treated with cryotherapy procedure after being diagnosed with precancerous cervical lesions at the time of the first screening were diagnosed with normal screening outcome. Moreover, 975(99.9%) of participants who were not suspected having invasive cervical cancer at the first screening, were diagnosed with normal screening outcome in a follow-up screening **(Table 2)**.

**Table 2:**
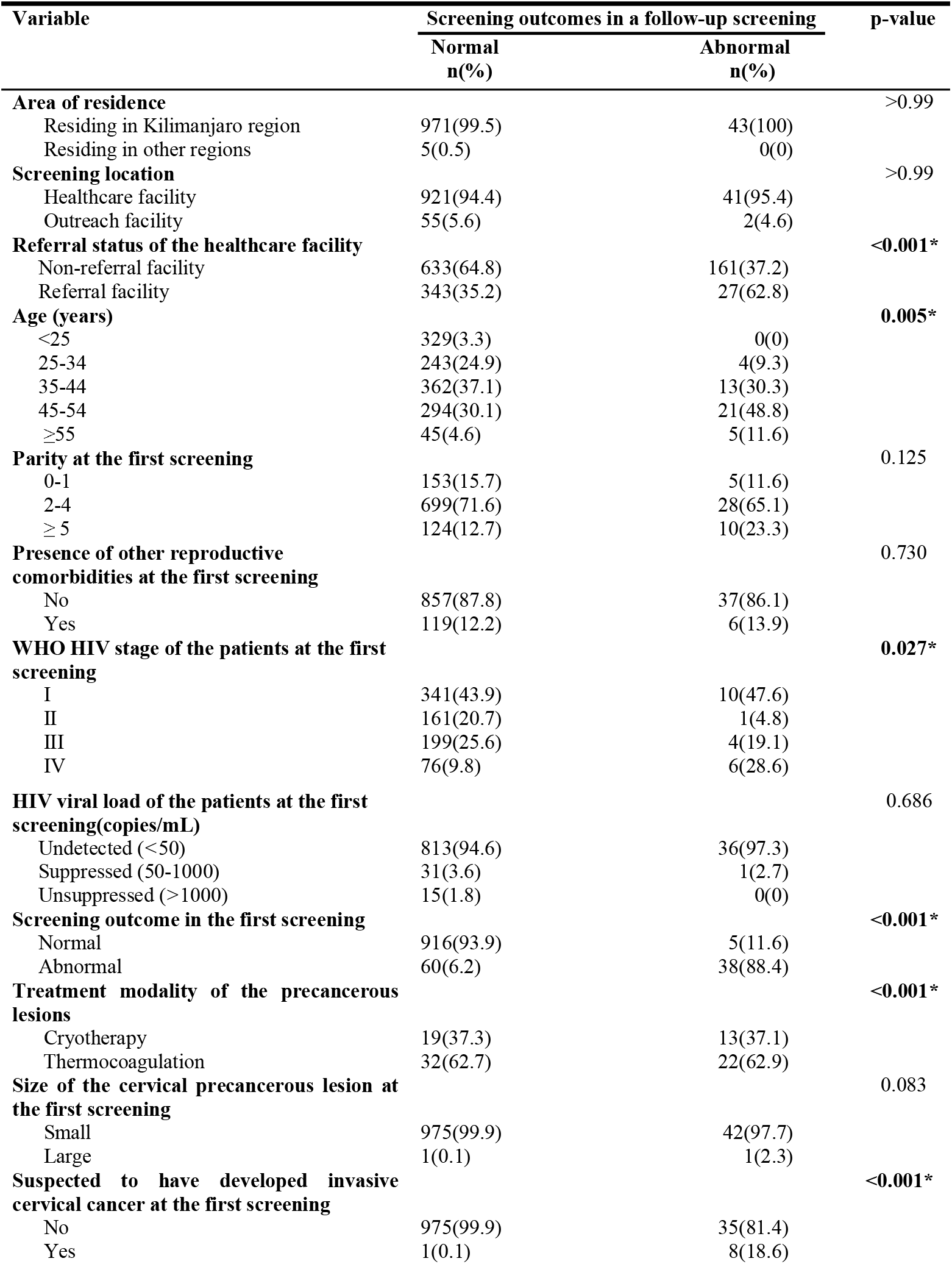
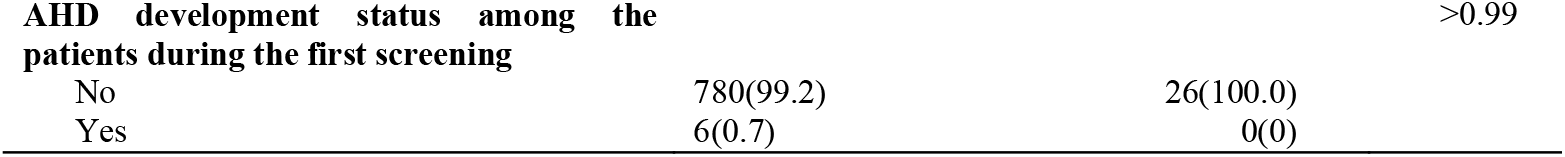
Screening outcomes in a follow-up cervical cancer screening among the study participants by their socio-demographic and clinical characteristics (n=1,019).

Moreover, in a multivariable analysis, after adjusting for the effect of other variables in the multivariable model, a statistically significant association was only found between the screening outcomes of the patients in a follow-up screening and their screening outcomes in their screening. Specifically, WLHIV who were diagnosed with abnormal screening outcomes in their first screening had 12.51 times higher odds of being yet again diagnosed with abnormal screening outcome in their follow-up screening compared to those who were diagnosed with normal screening outcomes in their first screening [AOR=12.51, 95%Cl: 8.92 – 37.64] **(Table 3)**.

**Table 3:** Multivariable analysis of the factors associated with the screening outcomes in a follow-up cervical cancer screening among WLHIV who had undertaken the first screening between 2019 and 2022 in Moshi Municipality (n=1,019).

| Variable | COR (95%CI) | p-value | AOR (95%CI) | p-value |
| --- | --- | --- | --- | --- |
| **Age (years)** |  |  |  |  |
| <25 | 1.00 |  | 1.00 |  |
| 25-34 | 0.15 (0.03 - 0.57) | 0.006 | 0.31 (0.01 - 7.37) | 0.473 |
| 35-44 | 0.32 (0.11 - 0.95) | 0.039 | 0.60 (0.05 - 8.02) | 0.701 |
| 45-54 | 0.64 (0.23 - 1.79) | 0.398 | 0.76 (0.06 - 9.69) | 0.831 |
| ≥55 | - |  | - |  |
| **Parity at the first screening** |  |  |  |  |
| 0-1 | 1.00 |  | 1.00 |  |
| 2-4 | 1.22 (0.47 - 3.23) | 0.680 | 0.62 (0.05 - 7.00) | 0.696 |
| ≥5 | 2.46 (0.82 - 7.41) | 0.107 | 0.91 (0.05 - 15.78) | 0.949 |
| **Presence of other reproductive comorbidities at the first screening** |  |  |  |  |
| No | 1.00 |  | 1.00 |  |
| Yes | 1.17 (0.48 - 2.82) | 0.731 | 2.31 (0.45 - 11.79) | 0.592 |
| **WHO HIV stage of the patients at the first screening** |  |  |  |  |
| I | 1.00 |  | 1.00 |  |
| II | 0.21 (0.03 - 1.67) | 0.141 | 0.28 (0.02 - 2.90) | 0.285 |
| III | 0.68 (0.21 - 2.22) | 0.528 | 0.68 (0.12 - 4.02) | 0.675 |
| IV | 2.69 (0.95 - 7.64) | 0.063 | 2.18 (0.46 - 10.29) | 0.326 |
| **HIV viral load of the patients at the first screening(copies/mL)** |  |  |  |  |
| Undetected (<50) | 1.00 |  | - |  |
| Suppressed (50-1000) | 0.73 (0.09 - 5.49) | 0.758 |  |  |
| Unsuppressed (>1000) | - |  |  |  |
| **Screening outcome in the first screening** |  |  |  |  |
| Normal | 1.00 |  | 1.00 |  |
| Abnormal | 16.03 (4.03 - 30.06) | >0.001 | 12.51(8.92 - 37.64) | >0.001* |
| **Size of the cervical precancerous lesion at the first screening** |  |  |  |  |
| Small | 1.00 |  | - |  |
| Large | 23.21 (1.43 - 37.56) | 0.027 |  |  |
| **AHD status** |  |  |  |  |
| No | 1.00 |  | - |  |
| Yes | 0.033 (0.02 - 0.05) | >0.001 |  |  |

## Discussion

The study revealed that 4.2% of WLHIV were diagnosed with abnormal screening outcome in the follow-up screening. A similar pattern was observed during their first screening whereby 4.0% of the screened WLHIV were diagnosed with abnormal screening outcome. These findings indicate a relatively low proportion of abnormal screening outcome among the participants in this study. This stands in sharp contrast to studies conducted in Kenya and Zimbabwe, which reported substantially higher proportions of abnormal screening outcome during a follow-up screening, 64.0% and 63.9%, respectively (8,15). Such disparities may be due to differences in population characteristics, screening programme implementation and/or diagnostic criteria across settings.

Moreover, the study also revealed that 88.4% of WLHIV who were diagnosed with abnormal screening outcome in the follow-up screening were also diagnosed with such screening outcomes during their first screening, despite having received treatment for cervical precancerous lesions shortly after the first diagnosis. This finding suggests a notably low regression of precancerous cervical lesions following treatment, raising concerns about the effectiveness of the treatment methods currently used used and the potential need for improved clinical management and closer monitoring of such cases (22).

On the other hand, this study found that 11.6% of participants who were diagnosed with abnormal screening outcome during the follow-up screening had initially received normal screening outcome during their first screening. This relatively high proportion may suggest the possibility of some of them having false-negative screening outcomes during their first screening. Such outcomes could have been attributed to the relatively low sensitivity and specificity of the screening modality used, inconsistencies in diagnostic interpretation, or variations in clinical judgement (23). These findings underscore the need to enhance the accuracy of cervical cancer screening through the use of more reliable diagnostic tools and standardized assessment criteria (24). In addition, the implementation of multiple testing approaches, either simultaneously or in parallel, could significantly increase the likelihood of correctly identifying the precancerous cervical lesions. Combining different screening methods such as VIA, HPV DNA test and pap smear test may offer improved diagnostic precision, particularly among high-risk WLHIV (25).

### Strength of the study

The study incorporated a wide range of potential factors affecting the screening outcomes in a follow-up screening, including both socio-demographic and clinical variables in the study. This comprehensive inclusion allowed for a more robust analysis and better understanding of the key factors associated with the screening outcomes in a follow-up cervical cancer screening among WLHIV.

### Limitations of the study

The registry from which data were obtained did not capture certain important demographic variables, such as educational attainment, marital status, employment status and ethnicity of the participants. This information could have provided a deeper understanding of even more social-demographic factors in influencing the screening outcomes in a follow-up cervical cancer screening

## Conclusion

A small proportion of WLHIV were diagnosed with abnormal screening outcome in a follow-up screening, majority of them had also been diagnosed with such a screening outcome in their first screening, despite being treated thereafter. This suggests a limited regression of precancerous cervical lesions despite prior treatment for such abnormalities.

Moreover, the only factor which was significantly associated with the screening outcomes in a follow-up cervical cancer screening was the screening outcomes in the first cervical cancer screening among the participants.

### Recommendations

The Ministry of Health should consider adopting multiple testing methods, such as combining VIA, Pap smear, and HPV DNA testing as standard practice for cervical cancer screening among WLHIV. This approach would help to reduce false-negative results which will lead to improved diagnostic accuracy and ensure early detection of precancerous cervical lesions.

### Ethical approval

Ethical approval for this study was obtained from the Ethical Review Committee of KCMC University, under clearance number PG 174/2024. In addition, permission to access data from the selected healthcare facilities in Moshi Municipality was granted by the respective authorities.

## Supporting information

Appendices.zip

## Data Availability

All data produced in the present study are available upon reasonable request to the authors

## Acknowledgements

We give our sincere gratitude to the Kilimanjaro Clinical Research Institute (KCRI) under the Network for Oncology Research in Africa (NORA) project for their financial support towards this work. We also thank the administration of the Moshi Municipality and the selected healthcare facilities for their invaluable cooperation during the data acquisition process.

## Authorship contribution statement

**Dickson Doto:** study conceptualization, design, data collection, analysis, report writing, and manuscript writing. **Laura Shirima:** data analysis, manuscript writing, review & editing, and supervision. **Blandina Mmbaga:** manuscript review & editing, mentorship and funding acquisition. **Innocent Mboya:** data analysis, manuscript review & editing, conceptualization and mentorship. **Innocent H. Peter Ughh:** manuscript writing, review & editing, **Alex Mremi**: manuscript writing, review & editing and mentorship. **Deogratius Kinoko:** manuscript writing, review & editing. **Patricia Swai:** data analysis, manuscript review & editing, conceptualization and supervision and **Bariki Mchome:** data analysis, manuscript review & editing, conceptualization and supervision.

## Declaration of Artificial Intelligence (AI) assistance

We acknowledge that AI tool (ChatGPT) was used solely to refine the clarity and readability of this work, without influencing its substantive ideas or content. All research activities – including data collection, analysis, results, and interpretation were entirely conducted by us and based exclusively on our original data.

---

**Figure 1:**
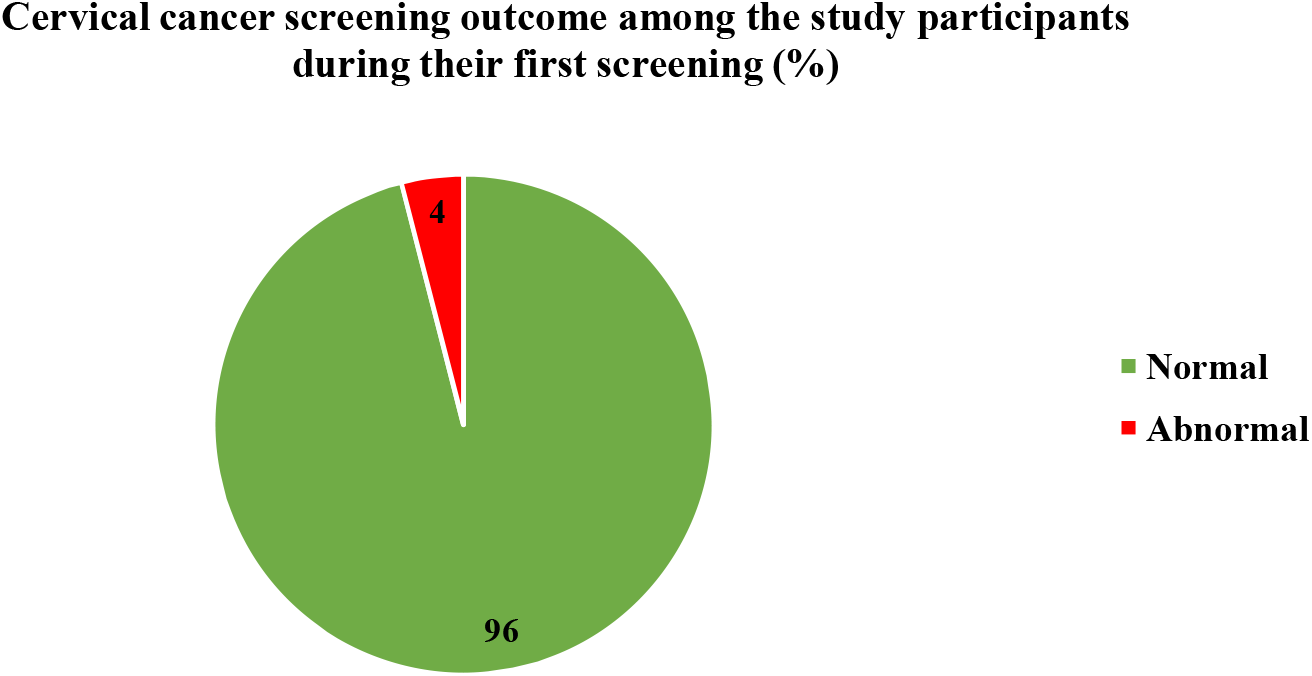
Cervical cancer screening outcome among the study participants during their first screening (n=3,076)

**Figure 2:**
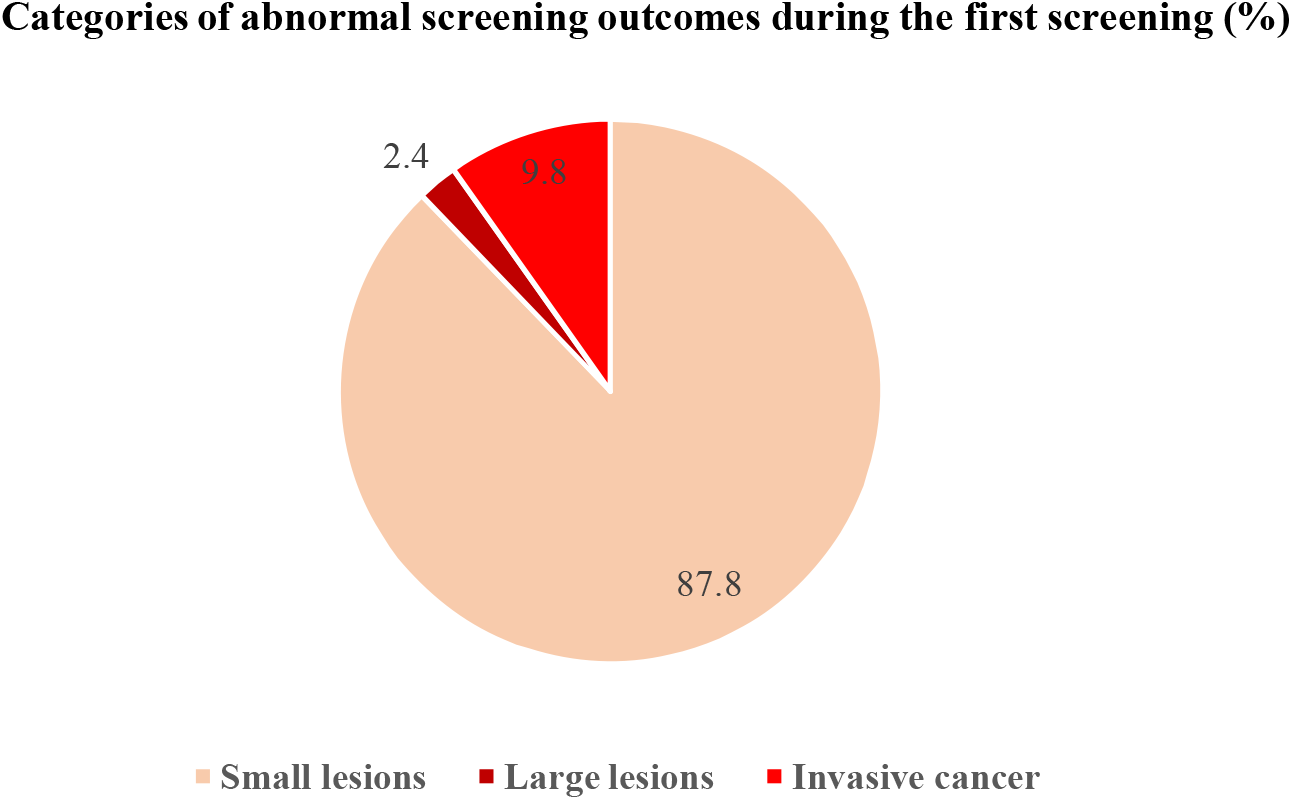
Categories of abnormal screening outcomes during the first screening (n=3,076)

**Figure 3:**
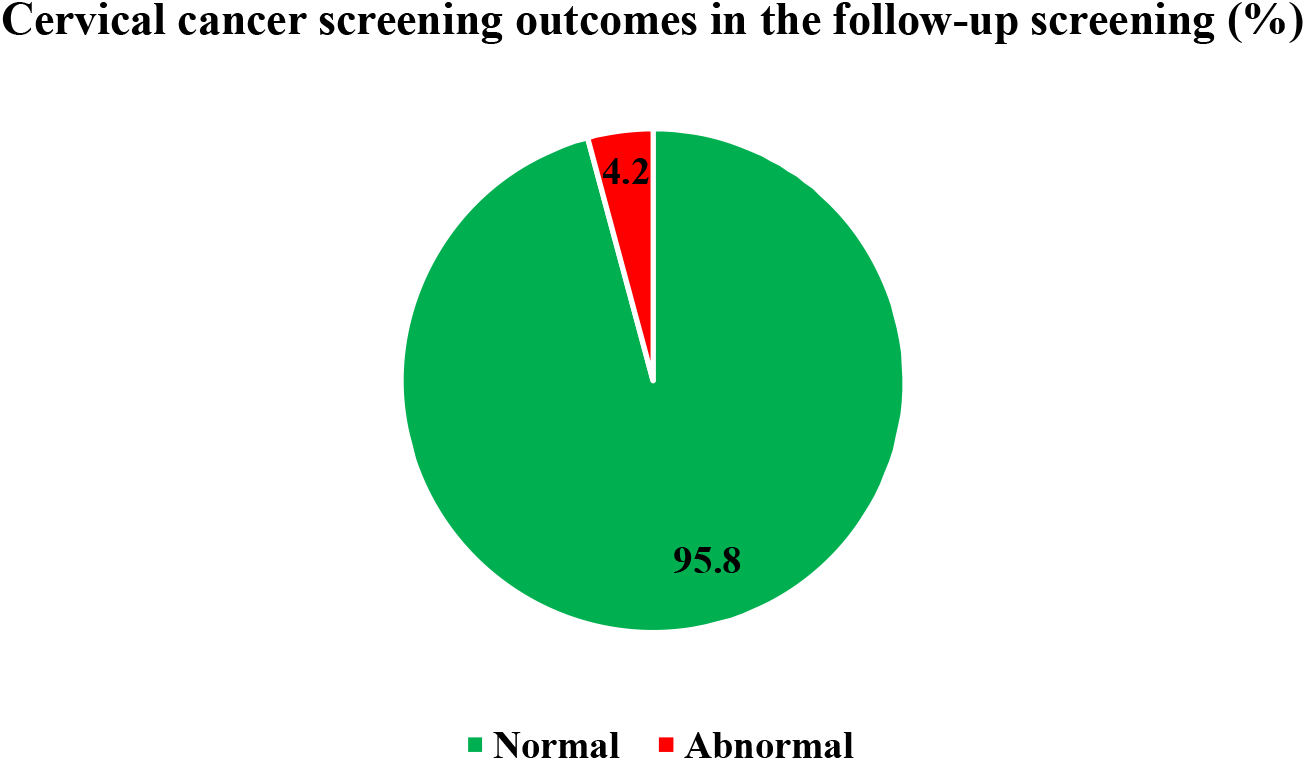
Cervical cancer screening outcomes in the follow-up screening (n=1,019)

