## Supplementary figures and images for "Screening Outcomes and Associated Factors in a Follow-up Cervical Cancer Screening among Women Living with HIV, 2019–2024: Evidence from Moshi Municipality"

### c15c8c7c-fdd7-482f-b88b-d0a2b4b8d524-0.jpg

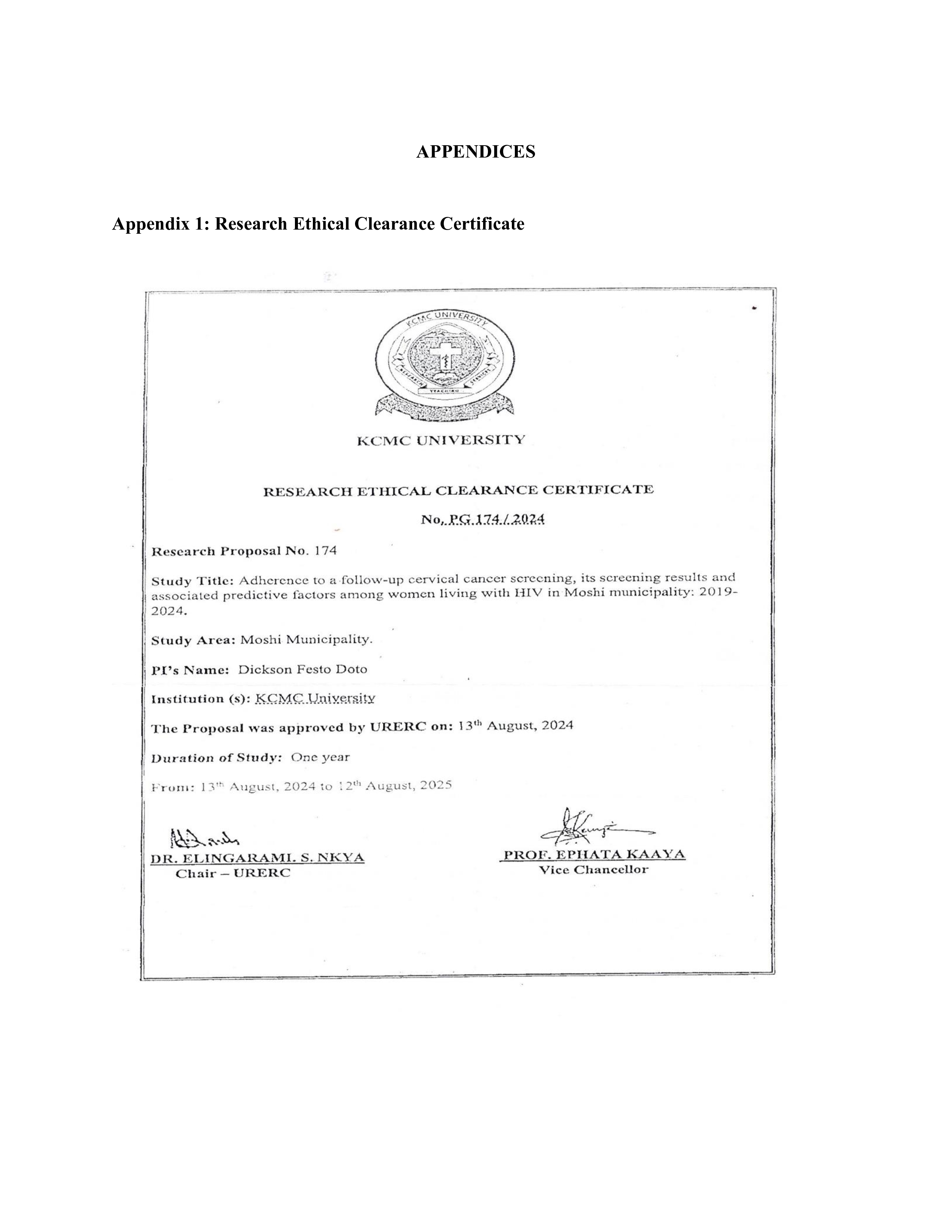

### c15c8c7c-fdd7-482f-b88b-d0a2b4b8d524-1.jpg

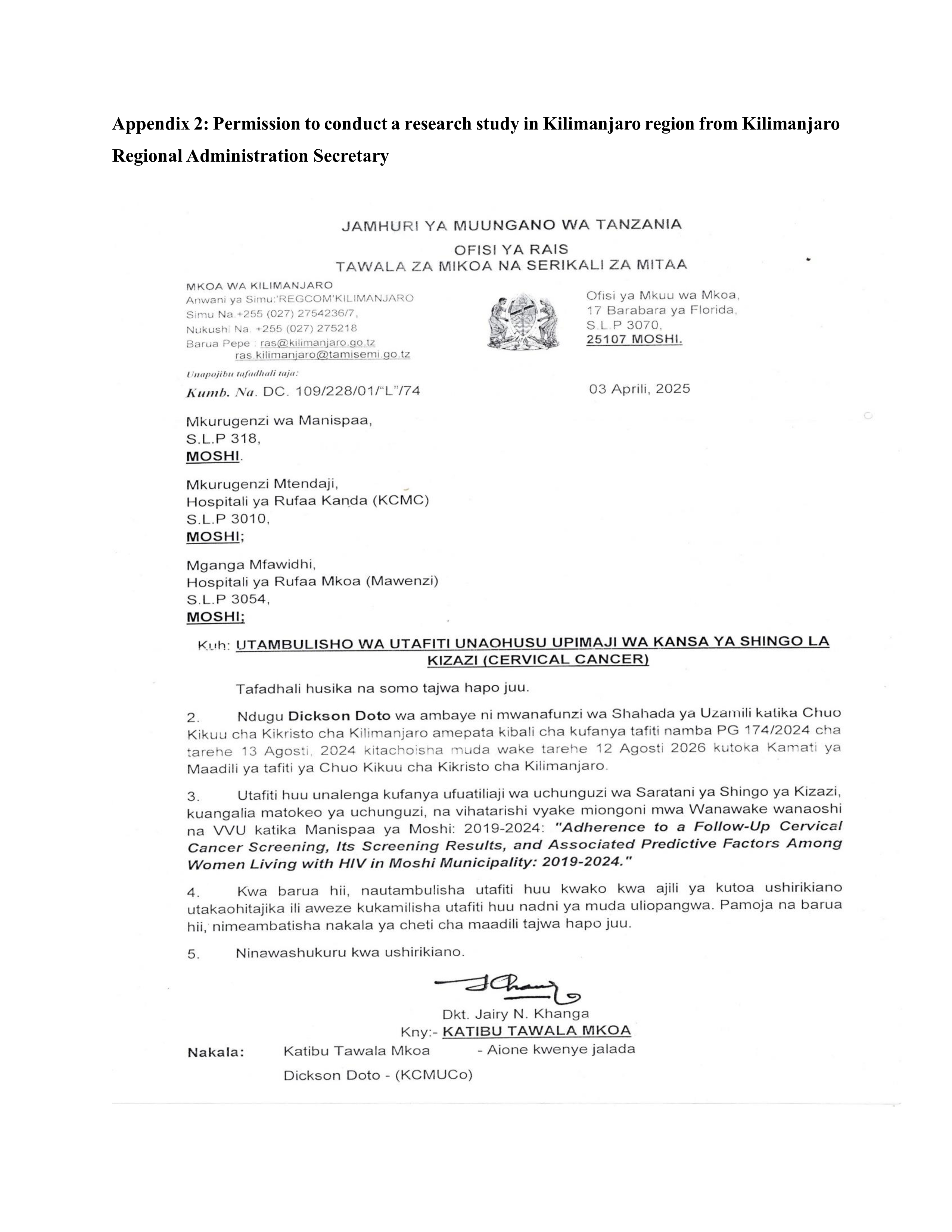

### c15c8c7c-fdd7-482f-b88b-d0a2b4b8d524-2.jpg

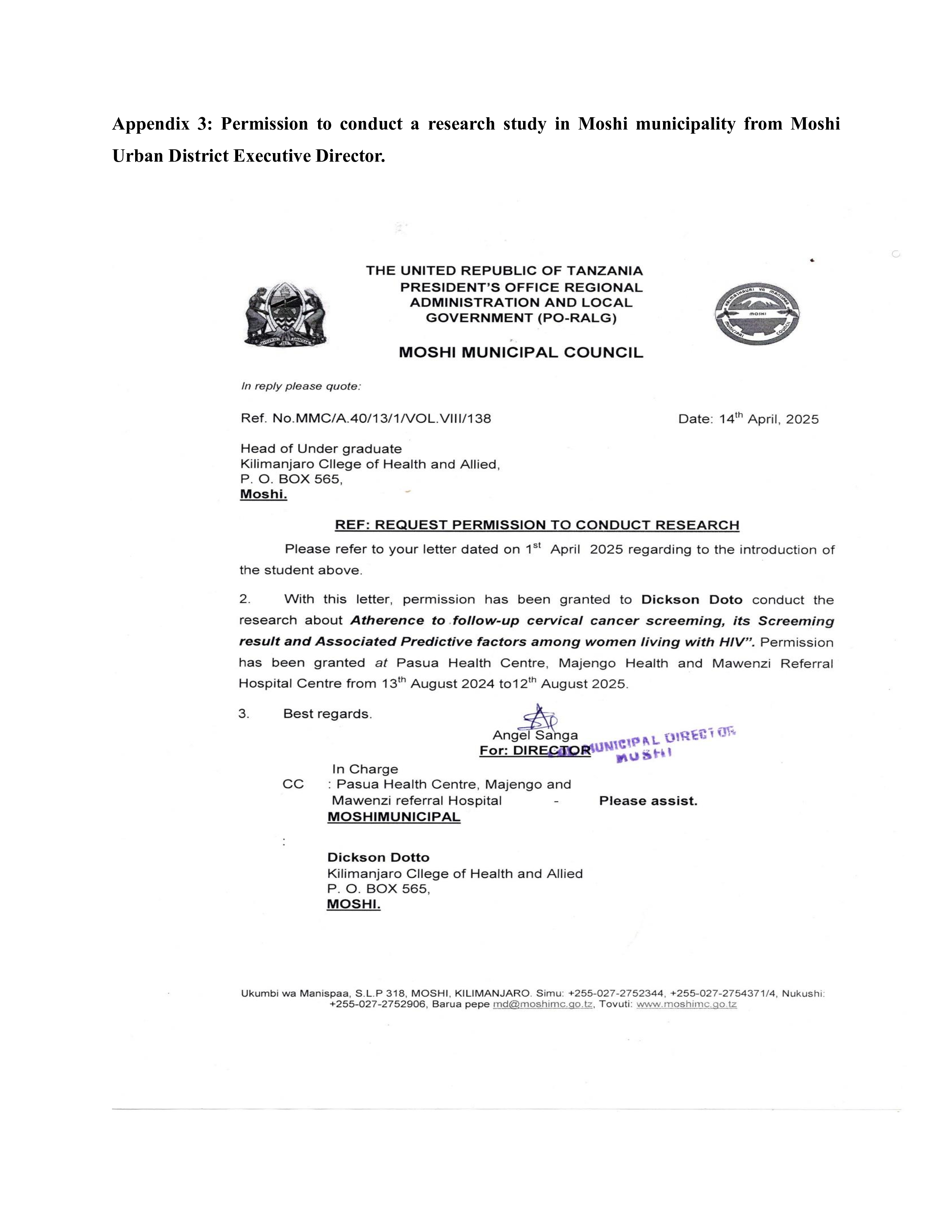

### c15c8c7c-fdd7-482f-b88b-d0a2b4b8d524-3.jpg

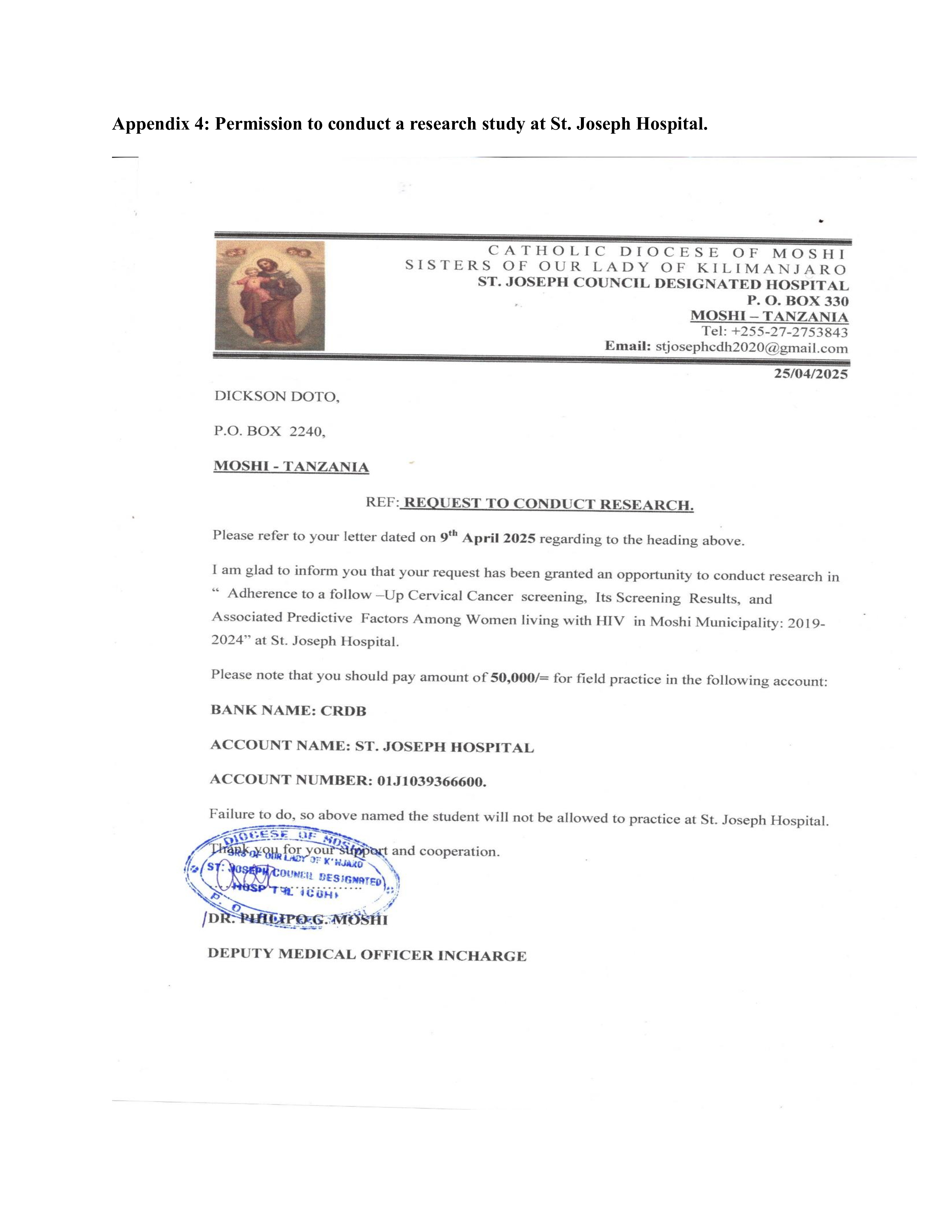
